# Spatial mapping of loco regional recurrences and disease related outcomes in breast cancer patients with Internal Mammary Node (IMN) positivity at presentation treated with a curative intent using moderately hypo-fractionated radiotherapy

**DOI:** 10.64898/2026.08.31.26361095

**Authors:** Divyani Chowdhury, Anurupa Mahata, Bhanu Vashistha, Santam Chakraborty, Sanjoy Chatterjee

**Author notes:** Corresponding author: Dr. Divyani Chowdhury, MBBS,MD, FRCR, Consultant, Department of Radiation Oncology, Tata Medical Center. Department of Radiation Oncology, All India Institute of Medical Sciences, Ansari Nagar, New Delhi 110029, India (Bhanu Vashistha). Dr. Sanjoy Chatterjee, MRCP, FRCR, FRCP, PGCE, FHEA, Senior consultant, Department of Radiation Oncology, Tata Medical Center,; Ms Anurupa Mahata, MSc DipRP, Senior Physicist, Tata Medical Center,; Dr. Bhanu Vashistha, MBBS,MD, FRCR, Fellow, Department of Radiation Oncology, Tata Medical Center,; Dr. Santam Chakraborty, MBBS,MD, Senior consultant, Department of Radiation Oncology, Tata Medical Center. **Institutional review board and ethical committee clearance no.:** EC/WV/TMC/26/25.

## Abstract

**Purpose**/**Objective:** There is paucity of data reporting outcomes of breast cancers with initial internal mammary nodal involvement and no visceral metastases, treated with curative hypofractionated radiotherapy. We report the outcomes from a tertiary centre alongside spatial patterns of recurrences in the above group

**Material/Methods:** For this retrospective cross-sectional study, consecutive patients contoured as per the ESTRO 2013 guidelines, treated between 2016-2022 were eligible if their diagnostic imaging demonstrated involvement of the internal mammary nodes. Radiotherapy (40 Gy/15#/3 weeks) was delivered to the residual breast / thoracic wall, SCF region corresponding to the ESTRO lymph node level 4 and internal mammary chain nodes. Residual IMN/ level 4 nodes received a boost of 10Gy/5#.

Spatial mapping of sites of recurrence at the local site and three nodal sites (axilla, SCF and IMN) was performed using deformable image registration.

Sites of recurrence at the local site and three nodal levels were contoured separately. Volumetric intersection of the recurrent gross tumour volume (GTV_recurrence) with treated clinical target volume (CTV) was calculated. Actuarial overall (OS), disease free survival (DFS) & cumulative incidence of local (LR), regional (RR) and loco-regional recurrence(LRR) were calculated using Kaplan Meier method. Univariate comparison of outcomes with or without residual disease was performed using the log rank test.

**Results:** The median age of the 61 eligible women was 49 years. 77% received neoadjuvant chemotherapy and the rest adjuvant chemotherapy. 82% patients had a mastectomy. Axillary lymph node dissection was done in 96.7%. Boosts to residual IMN and SCF nodes were delivered to 21(34.4%) and 2 (3.3%) respectively. Median follow up was 3.6 years. Out of the 61 patients, 42 patients were disease free with an estimated 3 year disease free survival of 75% (95% CI 64, 88%).

Spatial mapping of locoregional recurrence was possible in all but 1 patient with local (only) recurrence who was lost to follow-up after mammogram only. Among the patients with loco regional recurrence 1 had recurrence in local site + SCF +axilla, 3 had recurrence in the SCF+axilla, 2 in the SCF+IMN and 1 in the axilla+SCF+IMN. Only one patient had isolated axillary recurrence or isolated SCF recurrence. There were no IMN only recurrences. Among the 8 patients with nodal recurrence, a total of 27 individual GTV_recurrence were identified in the axilla(n=11), SCF(n=11) and IMN (n=5).

IMN recurrences showed complete or partial overlap with CTV. SCF recurrences were a mix with predominantly in-field recurrences while axillary recurrences occurred outside the treated volume.Four (6.6%) patients had Grade 2 lymphoedema as documented late side effect.

**Conclusion:** Aggressive treatment of IMN disease with adjuvant radiation is effective with good locoregional control. Systemic recurrences are common and may benefit from intensification strategies.

## Introduction

Breast cancer remains a significant global health issue, causing 670,000 deaths worldwide in 2022. In India, the incidence of breast cancer has seen a dramatic increase. According to GLOBOCAN 2020 data, breast cancer accounted for 13.5% of all cancer cases and 10.6% of all cancer deaths in India.(1–3). Advanced stage breast cancer, more commonly seen in women of lower socioeconomic status, is associated with worse prognosis and may represent an area of unmet clinical need (4).

IMN involvement upstages the AJCC TNM nodal staging of the disease to N2b (IMN only without axillary node) or N3b (IMN with axillary node)(5). Multiple randomized trials and meta-analyses show that locoregional recurrences outside the ipsilateral breast or chest wall have a worse prognosis, with IMN-metastasis breast cancer patients having a 5-year breast cancer-specific survival of 92.7, 77.4, and 66.0% in stage IIIA, IIIB, and IIIC diseases and the 5-year overall survival rates of 85.9, 72.1, and 58.7%, respectively.(6) Radical surgeries like extended mastectomy have shown no benefits for IMN-positive cases.(7) Current management involves a multimodal approach, including surgery for the breast and axillary nodes, systemic therapy, and radiotherapy targeting the breast and regional nodes, including the IMN region(8–10).

Elective internal mammary nodal radiotherapy has concerns about cardiac and pulmonary side effects(11). However recent meta analysis and large case series have reported better breast cancer specific survival and overall survival(12,13).

However, outcomes and recurrence patterns in patients with radiologically involved internal mammary nodes treated with curative intent radiotherapy remain sparsely reported.

## Materials and methods

### Study design

This retrospective cross-sectional study aimed to investigate locoregional recurrences and clinical outcomes in patients with IMN-positive breast cancer. This study was submitted to the Institutional Review Board for review. As this was a retrospective analysis of anonymized patient data, a formal ethical clearance waiver was granted.

### Aims and objectives

The primary goal was to perform spatial mapping of locoregional recurrences, identifying patterns and specific locations of recurrence in this high-risk cohort. The secondary objective focused on evaluating disease-related outcomes, including overall survival (OS) and disease-free survival (DFS), along with other relevant clinical metrics like local, nodal, distant and locoregional recurrence.

The primary objective was to map the spatial distribution of locoregional recurrences, like local, axillary, SCF and IMN recurrences to better understand recurrence patterns in IMN-positive cases. The secondary objective was to evaluate disease related outcomes by analyzing survival metrics.

### Inclusion and exclusion criteria

Patients included in the study were consecutive patients with IMN-positive breast cancer diagnosed on cross-sectional diagnostic imaging (contrast enhanced CT scan or ^18^FDG PET-CT) and treated between 2016 to 2022. Radiological evidence of IMN positivity was considered for IMN node size of 5 mm or higher or presence of uptake on FDG PET/CT scan.

### Surgery

All patients underwent definitive surgery to the breast and axilla. Breast surgery consisted of either mastectomy or breast conservation surgery, as clinically indicated. Axillary lymph node dissection (ALND) included complete dissection of axillary level I-III nodes.

### Radiotherapy planning

For patients receiving neoadjuvant chemotherapy followed by surgery and radiotherapy, the mean duration between surgery and radiotherapy was 44 days. On the other hand, for patients having surgery followed by adjuvant chemotherapy followed by radiotherapy, the mean duration between chemotherapy and the start of radiotherapy was 31 days. Patients were simulated in supine position with arms above the head using a breast board.

Intravenous contrast was used for better delineation of nodal regions based on vessel anatomy. Target volumes were drawn based on the ESTRO 2015 guidelines(14). For consistency with ESTRO nomenclature, the SCF volume referred to corresponds to ESTRO lymph node level 4.

### Physics planning

Patients were treated with Helical Tomotherapy using 6 MV photons.(15) Planning parameters included a pitch of 0.28, modulation factor of 2.5 with a field width of 2.5 mm to plan the three cases. Doses were optimized with the assistance of “dummy” to minimize dose dumping and low dose spillage. A dose of 40 Gy in 15 fractions over 3 weeks was prescribed. Any residual IMN received a sequential boost of 10 Gy in 5 fractions over 1 week.

### Data collection

Clinical, pathological and treatment related variables were retrospectively collected from institutional electronic medical records and radiotherapy databases. Molecular subtypes were classified based on ER, PR, HER2 and Ki-67 status. Luminal A tumours were defined as hormone receptor positive, Her2 negative tumours with Ki-67 <14%, while Luminal B tumours were hormone receptor positive tumours with HER2 positivity and/or Ki-67% >= 14%. HER2 enriched tumours were ER/PR negative and HER2-positive while triple-negative breast cancers were negative for ER,PR and HER2 (16).

### Image registration and spatial mapping

Individual sites of recurrence were delineated and labelled on the diagnostic imaging. Each distinct site of recurrence was considered an independent focus. Recurrences occurring in different nodal sites (axillary, supraclavicular, internal mammary) or at the local site (breast/chest wall) were contoured separately and designated as GTV_recurrence_axilla_1, GTV_recurrence_SCF_1, GTV_recurrence_IMN_1 and so on.

Using deformable registration, these diagnostic scans were fused with the initial planning CT images to evaluate overlap between the relapse sites and treatment volumes. Varian’s SmartAdapt feature which is a Deformable Image Registration (DIR) tool available with Eclipse treatment planning system was used for the purpose. It uses gradients in image intensity value to assist image registration. The process began with an initial rigid registration to align the images based on bony landmarks. Then the deformable registration was used which analyses voxel intensity patterns and anatomical contours to generate a deformable vector field (DVF), which is subsequently applied to adjust the anatomical structures accordingly, allowing for a more accurate match of the diagnostic images with the RT planning images. Acute and late side effects were graded according to the Common Terminology Criteria for Adverse Events, version 5.

### Statistical analysis

Statistical analysis was done by calculating the cumulative incidence of locoregional recurrence with a competing risk model and Kaplan Meier curves were used to calculate the overall survival (OS), disease free survival (DFS) and locoregional recurrence free survival (LRFS).

## Results

### Demographics and tumour details

The demographic information and tumour details has been summarised in Table 1.

**TABLE 1:**
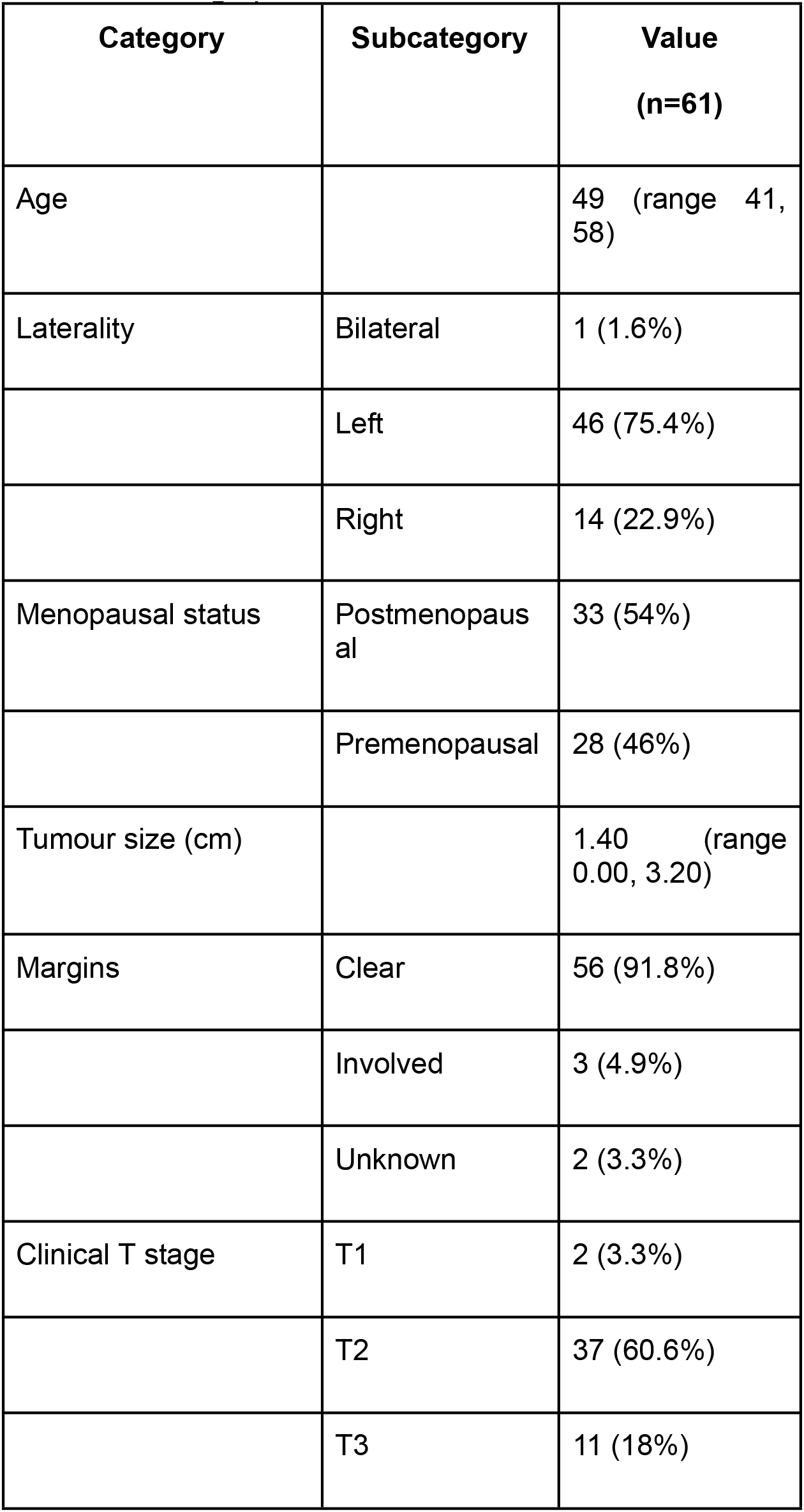
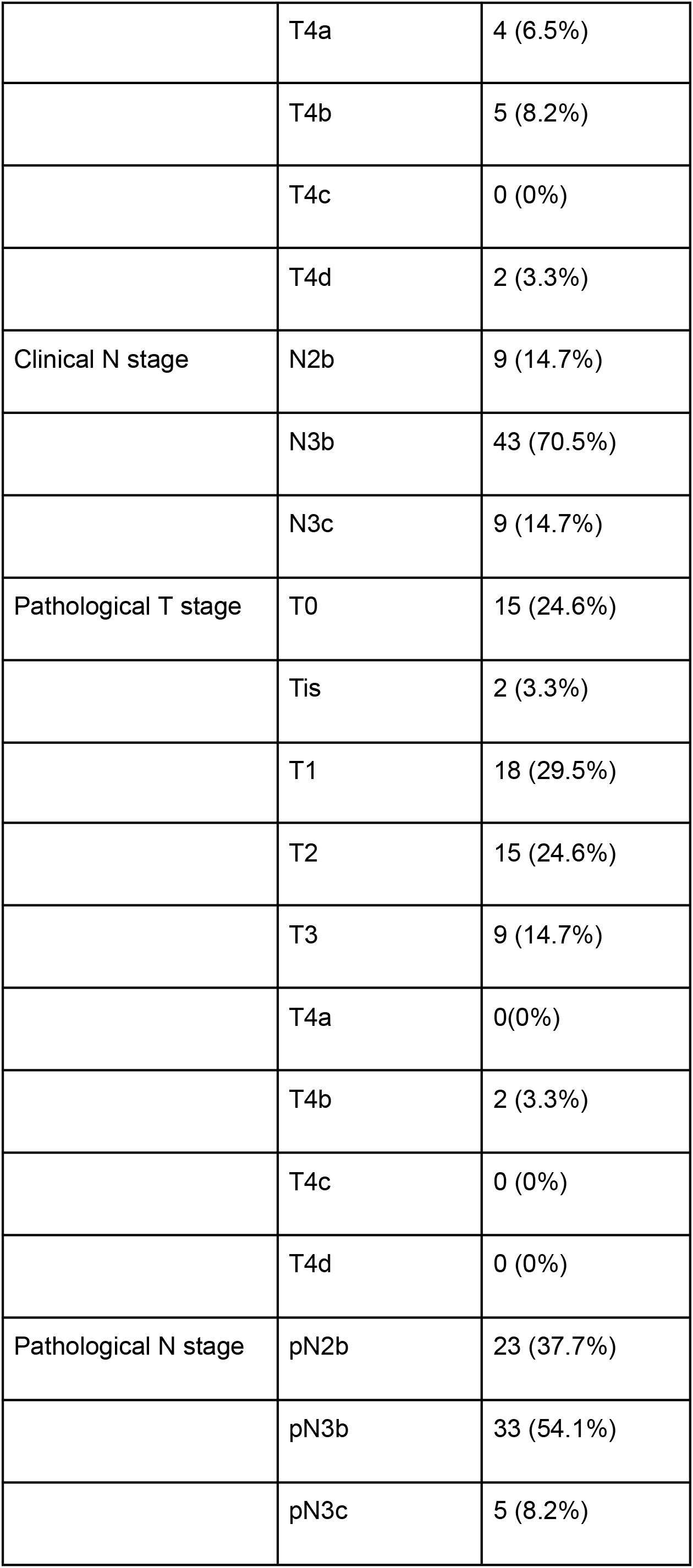
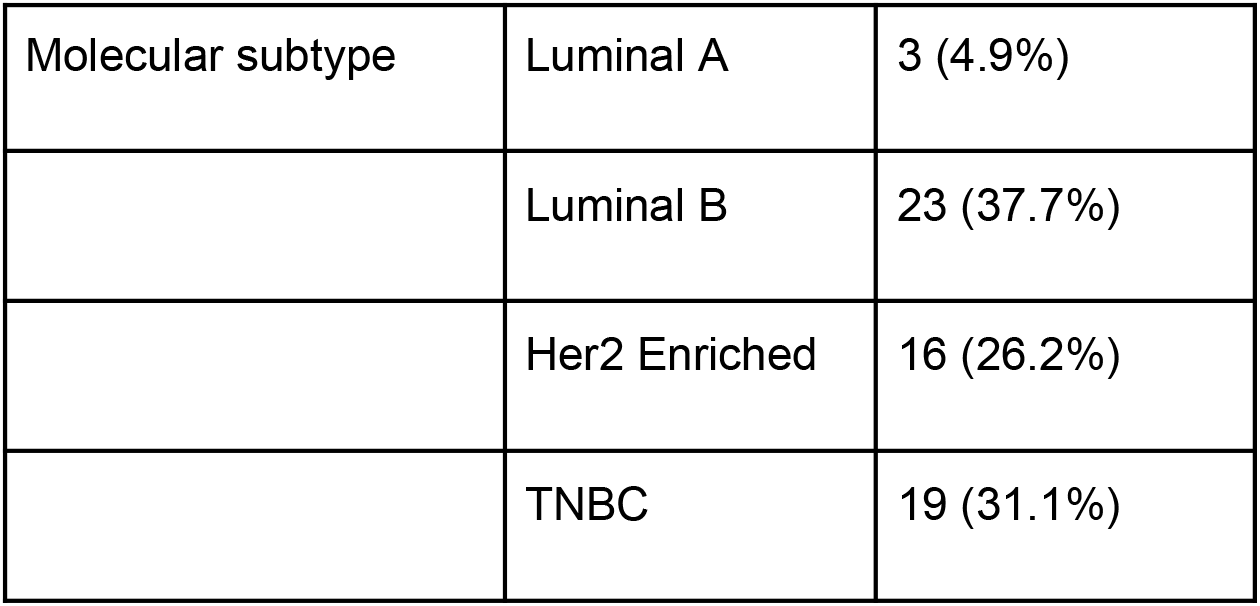
Demographics and tumour details.

| Category | Subcategory | Value<br>(n=61) |
| --- | --- | --- |
| Age |  | 49 (range 41, 58) |
| Laterality | Bilateral | 1 (1.6%) |
|  | Left | 46 (75.4%) |
|  | Right | 14 (22.9%) |
| Menopausal status | Postmenopausal | 33 (54%) |
|  | Premenopausal | 28 (46%) |
| Tumour size (cm) |  | 1.40 (range 0.00, 3.20) |
| Margins | Clear | 56 (91.8%) |
|  | Involved | 3 (4.9%) |
|  | Unknown | 2 (3.3%) |
| Clinical T stage | T1 | 2 (3.3%) |
|  | T2 | 37 (60.6%) |
|  | T3 | 11 (18%) |
|  | T4a | 4 (6.5%) |
|  | T4b | 5 (8.2%) |
|  | T4c | 0 (0%) |
|  | T4d | 2 (3.3%) |
| Clinical N stage | N2b | 9 (14.7%) |
|  | N3b | 43 (70.5%) |
|  | N3c | 9 (14.7%) |
| Pathological T stage | T0 | 15 (24.6%) |
|  | Tis | 2 (3.3%) |
|  | T1 | 18 (29.5%) |
|  | T2 | 15 (24.6%) |
|  | T3 | 9 (14.7%) |
|  | T4a | 0(0%) |
|  | T4b | 2 (3.3%) |
|  | T4c | 0 (0%) |
|  | T4d | 0 (0%) |
| Pathological N stage | pN2b | 23 (37.7%) |
|  | pN3b | 33 (54.1%) |
|  | pN3c | 5 (8.2%) |
| Molecular subtype | Luminal A | 3 (4.9%) |
|  | Luminal B | 23 (37.7%) |
|  | Her2 Enriched | 16 (26.2%) |
|  | TNBC | 19 (31.1%) |

All patients underwent definitive surgical management of the breast and axilla. Mastectomy was performed in 50/61 (82%) of patients, while 11/61(18%) underwent breast conservation surgery. ALND was performed in 96.7% of cases, and SLNB in the remaining 3.3%. The median number of nodes dissected was 21(IQR: 14-25), with a median number of positive nodes being 3 (IQR: 0-8).

All patients received some form of chemotherapy, neoadjuvant, adjuvant or both. The drugs used were 5-Fluorouracil, Epirubicin/Doxorubicin, Cyclophosphamide, Paclitaxel, Carboplatin and Capecitabine. NACT was administered to 77% of patients with the most common regimen being 4 cycles of dose dense FEC followed by paclitaxel. Sixteen patients received adjuvant chemotherapy. 54 patients completed 8 cycles with median number of chemotherapy cycles being 8. Fifty two out of sixty one(85.2%) patients received dose dense regimens. All hormone positive breast cancer patients received at least 5 years of tamoxifen or aromatase inhibitors. All 16 patients with Her2 positive disease received trastuzumab.

Two patients received pertuzumab along with trastuzumab. Eight of the TNBC patients who had residual disease after surgery received additional Capecitabine with the median number of Capecitabine being 8. None of the patients received immunotherapy.

Adjuvant radiotherapy was administered to all patients, delivered using tomotherapy. Radiotherapy volumes included chest wall/breast, supraclavicular fossa, and IMN. A sequential boost to the IMN was delivered to 34.4% of patients. The radiotherapy dose was 40 Gy in 15 fractions, with a boost dose of 10 Gy in 5 fractions.

### Outcomes

As of 1/11/24 with a median follow-up of 3.6 years, 49 patients were alive, while 12 had died. Local recurrence occurred in 2/61(3.3%) of patients, and nodal recurrence in 861(13.1%).7 patients had nodal recurrence in the supraclavicular fossa, 5 in the axilla and 3 in the IMN nodal regions. Distant metastases were observed in 16/61 (26.2%) patients, most commonly to the lungs (11/16,68.8%), followed by the brain (8/16,50%), bone (8/16,50%), and liver (5/16,31.3%). Figure 1 shows the upset chart depicting the patterns of locoregional recurrences. Patients with residual nodal disease at the time of radiotherapy had a numerically higher incidence of regional and distant recurrence compared to those with complete nodal response before radiotherapy as depicted in Table 2. The Kaplan Meier curves showing regional and distant recurrence of patients with and without nodal boost have been provided as Supplementary figures 1 and 2 respectively.

**FIGURE 1:**
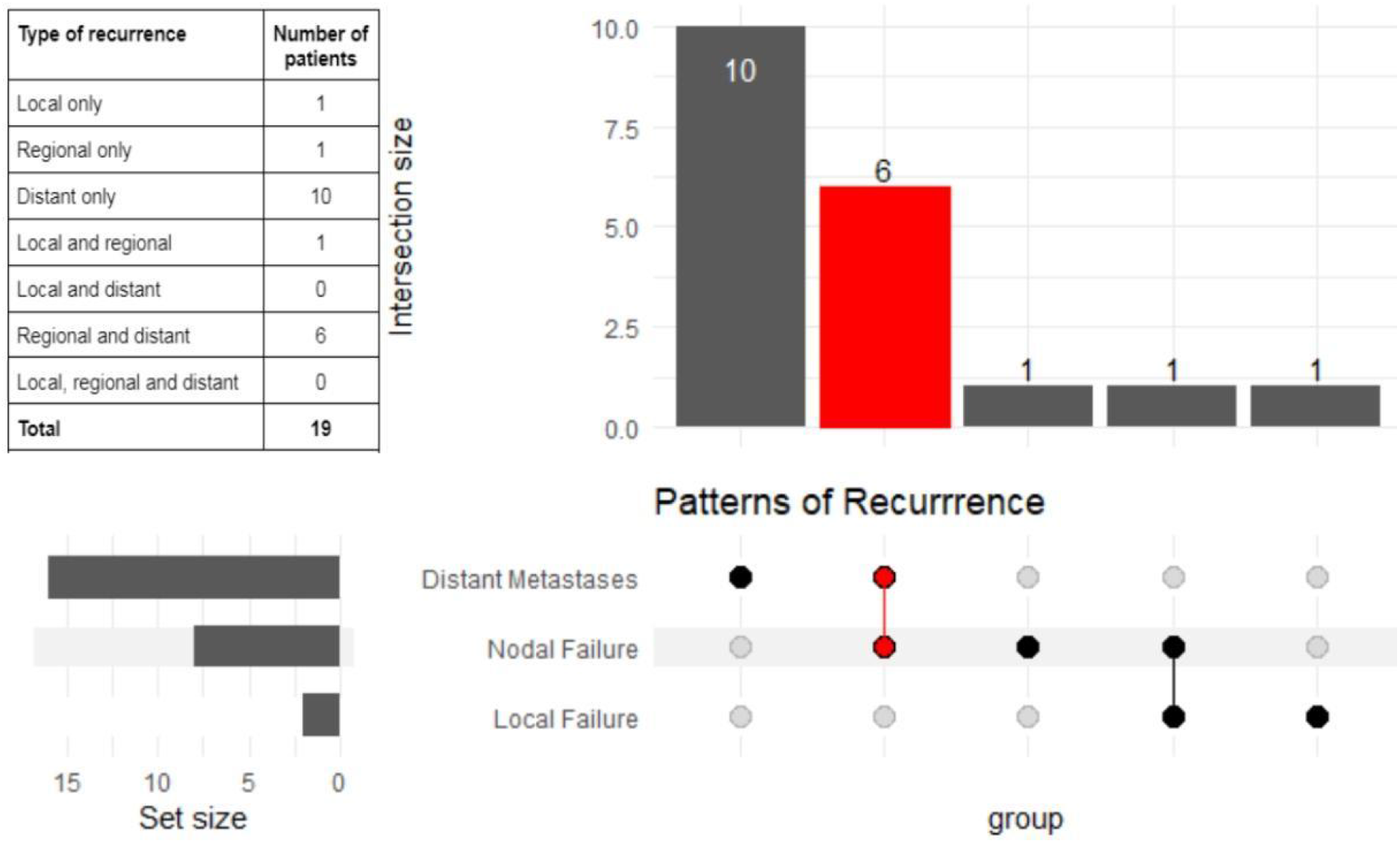
Figure showing offset chart showing type of recurrence

**TABLE 2:** Figure showing actuarial outcomes for eligible patients and comparison of outcomes between patients with or without residual nodal disease at the time of radiotherapy.

| Survival outcomes at 3 years | Overall<br>n=61 | Nodal residual<br>disease<br>n=23 | No nodal residual<br>n=38 |
| --- | --- | --- | --- |
| OS | 86%(77%, 96%) | 82% (64%, 100%) | 88% (78%, 100%) |
| DFS | 75% (64%, 88%) | 83% (71%, 96%) | 83% (71%, 96%) |
| Local recurrence<br>(n=2) | 3.6% (0%, 8.3%) | 5.3% (0%, 15%) | 7.1% (0%, 16%) |
| Nodal recurrence<br>(n=8) | 15% (4.0%, 25%) | 38% (3.2%, 60%) | 6.0% (0%, 14%) |
| Distant recurrence<br>(n=16) | 30% (15%, 42%) | 56% (17%, 77%) | 20% (5.5%, 33%) |
| Locoregional<br>recurrence(n= 9) | 17% (5.2%, 27%) | 38% (3.0%, 60%) | 8.6% (0%, 17%) |

### Spatial mapping

Among the 8 patients with nodal recurrence, a total of 27 discrete nodal recurrences were mapped and analysed using deformable image registration to determine volumetric overlap with the original radiotherapy fields as shown in Table 3. Figure 2 depicts the spatial distribution of SCF recurrences relative to the superior border of the subclavian vein, with circle sizes representing the volume of each recurrence and colors indicating the type: red for complete recurrences, green for partial, and blue for no overlap. More detailed information regarding the recurrences can be referred to in Supplementary Table 1.

**FIGURE 2:**
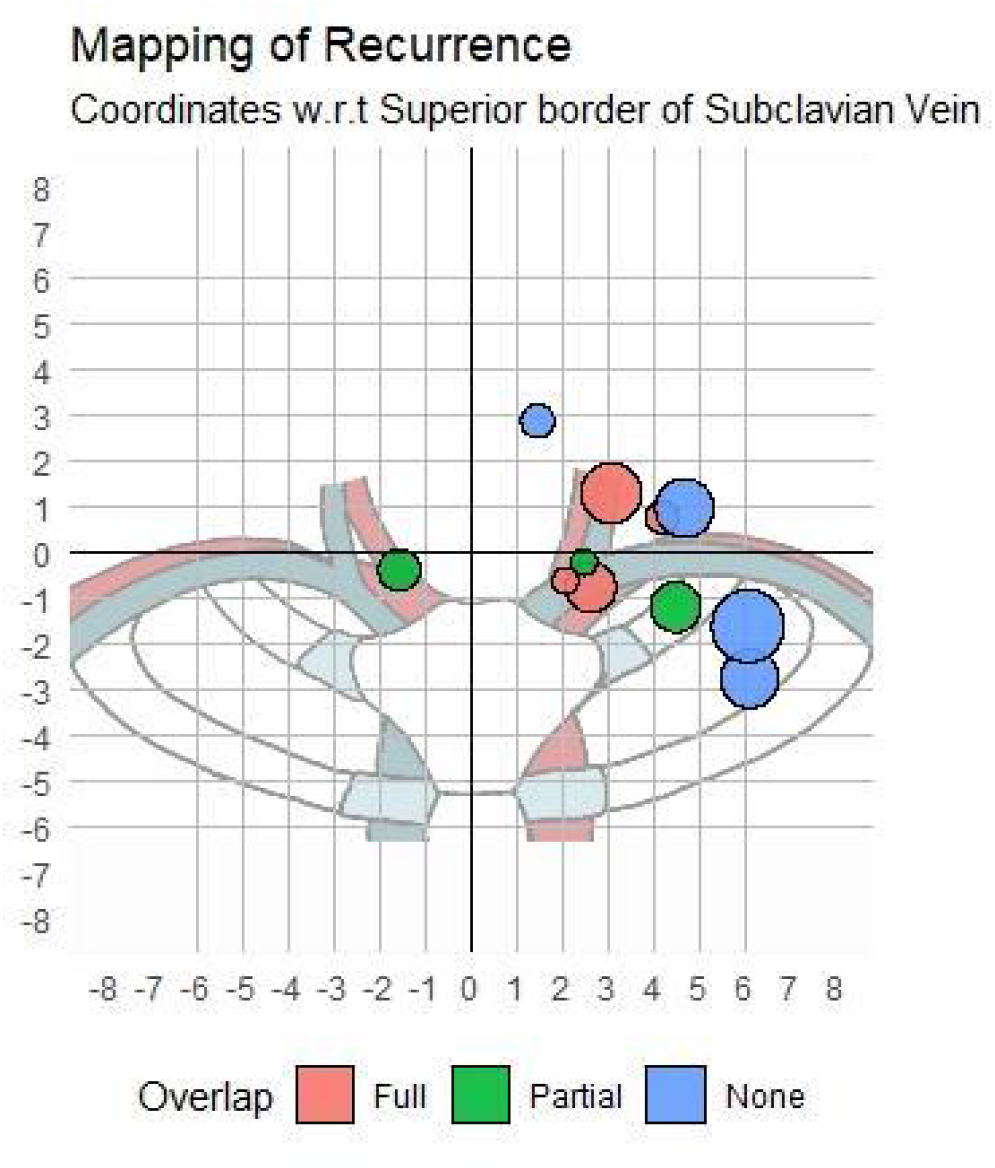
Figure showing spatial mapping of SCF recurrences with respect to the superior border of the subclavian vein. The numbers on the axis represent the distance in cm. Circle diameter represents the diameter of a volume-equivalent sphere derived from recurrence GTVs contoured and volume calculated in the eclipse treatment planning system after deformable registration. Color coding of the overlap indicates the overlap with the delineated target volume.

**TABLE 3:** Table showing sites along with the number of individual GTV_recurrence(in parenthesis) at each site for each patient with loco regional recurrences. A total of 9 patients had loco-regional recurrences.

|  | Local(n) | Axillary(n) | SCF(n) | IMN(n) |
| --- | --- | --- | --- | --- |
| Patient 1 |  | +(3) | +(2) |  |
| Patient 2 | +(1) |  |  |  |
| Patient 3 |  |  | +(1) |  |
| Patient 4 |  |  | +(2) | +(3) |
| Patient 5 | +(1) | +(3) | +(1) |  |
| Patient 6 |  | +(3) | +(2) | +(1) |
| Patient 7 |  | +(1) | +(2) |  |
| Patient 8 |  |  | +(1) | +(1) |
| Patient 9 |  | +(1) |  |  |
| **Total no of recurrences** | **2** | **11** | **11** | **5** |
| **Total number of patients with recurrences** | **2** | **5** | **7** | **3** |

IMN recurrences (n=5) showed high concordance with the irradiated CTV, with four showing complete overlap (100%) and one had partial overlap (56.5%). SCF recurrences (n=11) displayed variable overlap patterns. Five events showed complete overlap, two had partial overlap(14.3%) and four had no overlap. Axillary recurrences (n=11) were mostly outside the irradiated volume with only 2 showing partial overlap and nine occurring outside the radiotherapy field.

Side effect data were recorded for all patients included in the study (n = 61), based on their clinical notes available in the hospital electronic data records. All patients were followed up longitudinally, and the maximum grade of side effect was documented for each patient in accordance with the CTCAE. Lymphedema was reported in 9/61 (14.7%), primarily Grade 1 in 5/61 patients, followed by grade 2 and grade 3 in 3/61 and 1/61 patients respectively. Pneumonitis was observed in 2/61 (3.3%) patients, all Grade 1. Other late side effects, such as cardiac events (1/61,1.6%) were rare while rib fractures and brachial plexopathies were not observed. No cases of second malignancy were noted.

## Discussion

This study reports the clinical outcomes and spatial patterns of locoregional recurrence in a contemporary cohort of patients with imaging confirmed internal mammary node (IMN)-positive breast cancer treated with curative intent multimodality therapy, including moderately fractionated regional nodal radiotherapy. We observed good locoregional control, with low rates of recurrences despite the high risk nature of the cohort. Spatial mapping demonstrated that all IMN recurrences occurred within or partially within the irradiated clinical target volume, suggesting that recurrence in this region is more likely related to tumour biology. In contrast, most axillary recurrences occurred outside the treated target volume, while supraclavicular recurrences showed mixed in-field and out-of -field patterns.

Although locoregional control was favourable, distant metastasis remained the predominant mode of failure, particularly in patients with residual nodal disease after systemic therapy, highlighting the need for improved systemic treatment strategies.

The involvement of the internal mammary lymph nodes in breast cancer has long been associated with poor disease outcomes. Several studies indicate that positive IMN status correlates with decreased DFS (72%) and OS(81.2%) compared to those without IMN involvement (9,17), (18) Historical trials suggested a role for IMN dissection, but randomised trials have demonstrated that surgical removal or sentinel lymph node biopsy (SLNB) does not confer survival benefits(19)

Several key trials support the use of IMN directed RT reporting reduced recurrences and increased 5 year DFS (72%) and OS (77%)(20,21). Our results align with these findings, demonstrating that targeted IMN irradiation leads to a favourable local control with a low rate of IMN recurrence.

Despite its advantages, IMN radiotherapy presents significant planning challenges due to close proximity to critical structures such as the heart, lungs and brachial plexus. Ensuring adequate coverage while minimising dose to organs at risk requires complex planning. Radiotherapy for IMN in breast cancer can be delivered using various techniques, including 3DCRT, IMRT, VMAT, proton therapy, and electron-photon mixed beams, each offering varying levels of precision and organ sparing. Techniques like deep inspiration breath hold (DIBH) help reduce cardiac and lung doses, particularly for left-sided cancers. Advanced methods like proton therapy and helical tomotherapy provide superior dose conformity but are costly and less accessible. Our study utilised tomotherapy, ensuring precise dose delivery while maintaining accepted side effect profiles. According to Yang et al, dose escalation to IMN (>=63.6Gy) was associated with improved DFS in patients with IMN size >=1cm, reinforcing the importance of individualised treatment planning.(18) The balance between optimal tumor coverage and organ at risk sparing remains a critical point.

While IMN directed RT provides clear oncologic benefits, it also introduces potential side effects, including pulmonary, cardiac complications, rib fracture and brachial plexopathy. Our findings revealed a low incidence of grade 2 or higher side effects, with pneumonitis observed in only 3.3% of patients and a single reported cardiac event (1.6%), consistent with published literature. These results suggest that modern RT techniques can effectively mitigate severe side effects while maintaining treatment efficacy. However, a limitation of the present study is that side effect data were collected retrospectively from available clinical and electronic medical records. Although every effort was made to capture and verify side effect outcomes for all included patients, the retrospective nature of data collection may introduce the possibility of under-reporting or incomplete documentation when compared with prospective studies. Therefore long-term, prospective follow-up is necessary to assess late toxic effects, particularly given the risk of cardiac morbidity associated with left-sided IMN irradiation.

Understanding recurrence patterns in IMN-positive patients is crucial for optimizing treatment strategies. A key strength of this study is the use of deformable image registration in which cross sectional images were reviewed to identify relapses, with diagnostic scans fused to planning CTs using Varian’s SmartAdapt for Deformable Image Registration. This feature, involving initial rigid registration followed by deformable registration based on voxel intensity and anatomical contours to accurately map relapse sites, has been validated in several studies as a powerful tool with sufficient accuracy for radiotherapy volume delineation (22–24).This approach allowed a more precise spatial correlation between relapse location and previously treated volumes improving the robustness of the study.

Our data indicate that while local and nodal recurrences are relatively low, systemic recurrence remains a major concern, with 16 patients experiencing distant recurrences. The most common sites of metastasis being the lungs (11/16, 68.8%), liver (5/16, 31.3%), brain (8/16, 50.0%), and bone (8/16, 50.0%). These findings emphasize the need for effective systemic therapy in addition to locoregional treatment.

The majority of our cohort received neoadjuvant chemotherapy and targeted therapy. However none of the patients were given immunotherapy. The relatively high rate of distant recurrences suggests that future studies should explore the role of intensified systemic treatments in improving survival outcomes for IMN-positive breast cancer patients. Similarly, Joo et al. reported that 85% of their cohort received systemic therapy, including anthracycline-based chemotherapy, taxanes, and targeted therapy for HER2-positive patients(21). Their study demonstrated that patients who received comprehensive systemic treatment in addition to RT had significantly improved survival outcomes compared to those who did not. The relatively high rate of distant recurrences in our study suggests that future research should focus on optimizing systemic treatment strategies in IMN-positive patients to improve survival outcomes.

A detailed analysis of nine patients with local and regional relapse showed a heterogeneous pattern of recurrence across different anatomical regions. IMN relapses were most commonly associated with a high degree of overlap (100%) with both the clinical target volume (CTV) and planning target volume (PTV), indicating that despite adequate radiation coverage, local IMN disease progression was due to underlying tumour radioresistance.. Conversely, axillary and supraclavicular (SCF) relapses displayed variable overlap patterns, with some cases showing complete coverage and others exhibiting no overlap, suggesting under-treatment or disease progression beyond planned radiation fields. Some SCF and IMN relapses exhibited partial overlap with CTV/PTV, raising questions about dose sufficiency and potential areas of geographic miss. These findings highlight the complexities of treatment planning and emphasize the need for further research into adaptive radiotherapy strategies, potential dose escalation in high-risk areas, and improved imaging techniques to enhance precision in recurrence detection and management. A limitation of this study is that recurrence mapping was based on volumetric overlap with target volumes and detailed dosimetric assessment of delivered dose and recurrence overlap could not be performed due to the retrospective nature of the dataset.

A very important finding was that both regional and locoregional recurrences were seen to be numerically more in patients who had residual IMN nodes even after these patients received boost to the residual nodes.However, due to the small sample size statistical significance could not be established. In addition, the boost dose administered to residual disease reflected institutional practice during the study period, and the optimal dose escalation strategy for residual nodal disease remains uncertain. Future research efforts need to be directed towards intensification of both local and systemic therapy in this subgroup of patients.

The present study has some limitations. As a retrospective single-centre analysis, it is subject to inherent selection bias and treatment heterogeneity. The relatively small sample size limits the statistical power of the study making robust subgroup analysis difficult.

Furthermore, toxicity assessment was retrospective and based on documentation in electronic medical records, introducing the potential for under-reporting and reporting bias compared with prospectively collected toxicity data. In addition to this, there remains no consensus regarding the optimal radiotherapy dose for residual nodal disease in breast cancer. The boost dose used in this study reflected institutional practice during the study period, and whether higher doses may improve local control without increasing toxicity warrants prospective evaluation. Finally, the retrospective nature of the study precluded detailed dosimetric assessment of delivered dose at sites of recurrence, limiting our ability to correlate dose distribution with patterns of failure.

## Conclusion

Use of adjuvant IMN radiotherapy in patients with involved IMN at presentation results in encouraging disease control and support the use of IMN irradiation, suggesting favorable local outcomes with manageable side effects. However, systemic recurrence remains a major challenge, underscoring the importance of comprehensive multimodal treatment strategies. Future research should focus on refining radiation dose optimization and improving systemic therapy to enhance survival outcomes for IMN-positive patients.

*Artificial intelligence (ChatGPT, OpenAI) was used solely to improve language, grammar and readability of the manuscript. The authors have reviewed and approved all revisions and take full responsibility for the final content*.

## Data Availability

All data produced in the present study are available upon reasonable request to the authors

## Abbreviations

ALND: Axillary Lymph Node Dissection
CT: Computed Tomography
CTCAE: Common Terminology Criteria for Adverse Events
CTV: Clinical Target Volume
DCIS: Ductal Carcinoma In Situ
DFS: Disease-Free Survival
DIR: Deformable Image Registration
DVF: Deformable Vector Field
ER: Estrogen Receptor
FDG PET-CT: Fluorodeoxyglucose Positron Emission Tomography - Computed Tomography
GTV: Gross Tumor Volume
HER2: Human Epidermal Growth Factor Receptor 2
IMN: Internal Mammary Node
LVI: Lymphovascular Invasion
LR: Local Recurrence
LRC: Loco-regional Recurrence
NACT: Neoadjuvant Chemotherapy
OS: Overall Survival
PR: Progesterone Receptor
PTV: Planning Target Volume
RR: Regional recurrence
RT: Radiotherapy
SCF: Supraclavicular Fossa
SLNB: Sentinel Lymph Node Biopsy
TNBC: Triple-Negative Breast Cancer
VMAT: Volumetric Modulated Arc Therapy

