## Supplementary Material for "Spatial mapping of loco regional recurrences and disease related outcomes in breast cancer patients with Internal Mammary Node (IMN) positivity at presentation treated with a curative intent using moderately hypo-fractionated radiotherapy"

SUPPLEMENTARY DATA FILE

SUPPLEMENTARY TABLE 1:

Table showing overlap of individual recurrences with CTV for all patients with nodal recurrences.

| Patient No. | Local relapse | Overlap with CTV | Axillary relapse | Overlap with CTV | SCF relapse | Overlap with CTV | IMN relapse | Overlap with CTV |
| --- | --- | --- | --- | --- | --- | --- | --- | --- |
| 1 | - | - | 1 | Partial  (33.33%) | 1 | Complete  (100%) | - | - |
|  |  |  | 2 | No  (0%) |  |  |  |  |
|  |  |  |  |  | 2 | Complete  (100%) |  |  |
|  |  |  | 3 | no  (0%) |  |  |  |  |
| 2 | 1 | complete | - | - | - | - | - | - |
| 3 | - | - | - | - | 1 | No  (0%) | - | - |
| 4 | - | - | - | - | 1 | Complete  (100%) | 1 | Complete  (100%) |
|  |  |  |  |  |  |  | 2 | Complete  (100%) |
|  |  |  |  |  | 2 | No  (0%) |  |  |
|  |  |  |  |  |  |  | 3 | Complete  (100%) |
| 5 | 1 | complete | 1 | Partial  0.9 (60%) | 1 | Partial  (14.29%) | - | - |
|  |  |  | 2 | No  0 (0%) |  |  |  |  |
|  |  |  | 3 | No  0 (0%) |  |  |  |  |
| 6 | - | - | 1 | No  (0%) | 1 | No  (0%) | 1 | Partial  (56.52%) |
|  |  |  | 2 | No  (0%) |  |  |  |  |
|  |  |  |  |  | 2 | No  0 (0%) |  |  |
|  |  |  | 3 | No  (0%) |  |  |  |  |
| 7 | - | - | 1 | Complete  (100%) | 1 | Complete  (100%) | - | - |
|  |  |  |  |  | 2 | Complete  (100%) |  |  |
| 8 | - | - | - | - | 1 | Partial  (75%) | 1 | Complete  (100%) |
| 9 | - | - | 1 | No  (0%) | - | - | - | - |

SUPPLEMENTARY TABLE 2:

Table showing dosimetric constraints for radiotherapy planning.

Supplementary Table 2A. Whole Breast (BCS) + SCF + IMN with Simultaneous Integrated Boost (40 Gy + 48 Gy in 15 fractions)

| **Structure** | **Parameter** | **Absolute dose** | **Optimal** | **Mandatory** |
| --- | --- | --- | --- | --- |
| Boost Target Volume (BTV) | V95% of 48 Gy | 45.6 Gy | ≥95% | ≥90% |
|  | V107% of 48 Gy | 51.4 Gy | – | <2% |
|  | Maximum dose (D0.5cc) |  | – | ≤52.8 Gy (110% of 48 Gy) |
| PTV WB – BTV | V95% of 40 Gy | 38 Gy | ≥95% | ≥90% |
|  | V100% of 48 Gy | 48 Gy | ≤5% | – |
| PTV WB – BTV +1 cm | V107% of 40 Gy | 42.8 Gy | ≤5% | – |
|  | V110% of 40 Gy | 44 Gy | ≤2 cc | – |
| Individual nodal PTV | V90% of 40 Gy | 36 Gy | ≥90% | – |
|  | V80% of 40 Gy | 32 Gy | – | ≥80% |
|  | V107% of 40 Gy | 42.8 Gy | – | ≤2% |
|  | Maximum dose (D0.5cc) |  | – | ≤44 Gy |
| Ipsilateral lung | Mean dose |  | ≤13 Gy | V17Gy ≤35% |
| Contralateral lung | Mean dose |  | ≤4 Gy | – |
| Heart | Mean dose |  | ≤4 Gy | ≤6 Gy  V17Gy ≤10% |
| Contralateral breast | Mean dose |  | <1 Gy (<40 years of age); <1.5 Gy (≥40 years of age) | ≤3.5 Gy |
| Oesophagus | Mean dose |  | ≤11 Gy  V17Gy ≤15% | – |
| PRV brachial plexus | Maximum dose |  | – | ≤40 Gy |

Supplementary Table 2B. Chest Wall (MRM) + SCF + IMN (40 Gy in 15 fractions)

| **Structure** | **Parameter** | **Absolute dose** | **Objective** | **Constraint** |
| --- | --- | --- | --- | --- |
| PTV WB | V95% of 40 Gy | 38 Gy | ≥95% | ≥90% |
|  | V105% of 40 Gy | 42 Gy | ≤5% | ≤7% |
|  | V107% of 40 Gy | 42.8 Gy | – | ≤2% |
|  | Maximum dose (D0.5cc) |  | – | ≤44 Gy (110%) |
| Individual nodal PTV | V90% of 40 Gy | 36 Gy | ≥80% | – |
|  | V80% of 40 Gy | 32 Gy | – | ≥90% |
|  | V107% of 40 Gy | 42.8 Gy | – | ≤2% |
|  | Maximum dose (D0.5cc) |  | – | ≤44 Gy |
| Ipsilateral lung | Mean dose |  | ≤13 Gy | V17Gy ≤35% |
| Contralateral lung | Mean dose |  | ≤4 Gy | V2.5Gy ≤15% |
| Heart | Mean dose |  | ≤4 Gy | ≤6 Gy  V17Gy ≤10% |
| Contralateral breast | Mean dose |  | <1 Gy (<40years of age); <1.5 Gy (≥40years of age) | ≤3.5 Gy |
| Oesophagus | Mean dose |  | ≤11 Gy  V17Gy ≤15% | – |
| PRV brachial plexus | Maximum dose |  | – | ≤40 Gy |

SUPPLEMENTARY FIGURE 1:

KM curves showing regional/nodal recurrence of patients with and without nodal boost. Patients receiving a nodal boost had gross residual disease at the time of radiotherapy, whereas patients not receiving a boost had no residual nodal disease and received elective nodal irradiation only.

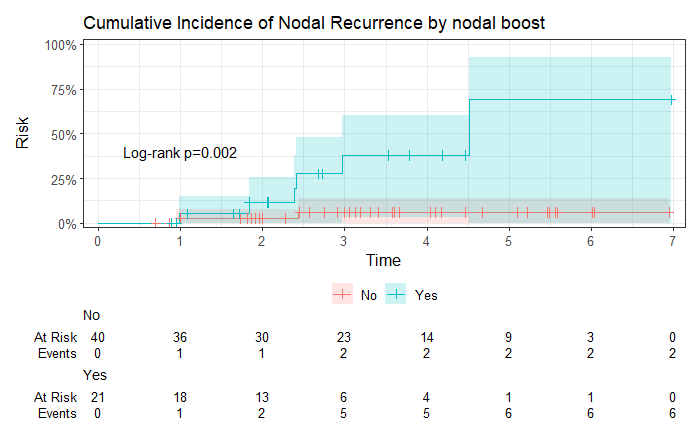

SUPPLEMENTARY FIGURE 2:

KM curves showing distant metastasis of patients with and without nodal boost. Patients receiving a nodal boost had gross residual disease at the time of radiotherapy, whereas patients not receiving a boost had no residual nodal disease and received elective nodal irradiation only.

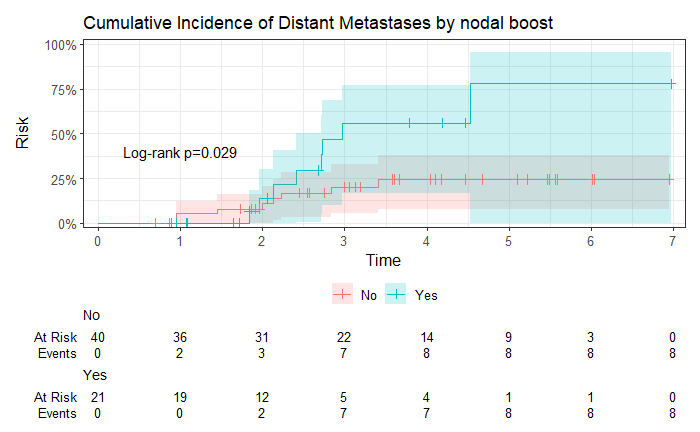
